# Longitudinal Changes in the Carotid Arteries of Head and Neck Cancer Patients Following Radiation Therapy: Results from a Prospective Serial Imaging Biomarker Characterization Study

**DOI:** 10.1101/2023.09.18.23295583

**Authors:** MD Anderson Head and Neck Radiation Oncology and Cardiovascular Working Group, Efstratios Koutroumpakis, Abdallah Sherif Radwan Mohamed, Peter Chaftari, David I. Rosenthal, Dorothy Gujral, Christopher Nutting, Peter Kim, Roland Bassett, Clifton D. Fuller, Elie Mouhayar

**Author notes:** Addresses for correspondence: 1. Clifton D. Fuller, MD Professor Division of Radiation Oncology The University of Texas MD Anderson Cancer Center 1515 Holcombe Blvd, Houston, TX 77030 Email address 2. Elie Mouhayar, MD Professor of Medicine Department of Cardiology, Division of Internal Medicine The University of Texas MD Anderson Cancer Center 1515 Holcombe Blvd # 1451, Houston, TX 77030. The 2 senior authors contributed equally (Clifton D. Fuller, Elie Mouhayar).

## Abstract

**INTRODUCTION:** We prospectively evaluated morphologic and functional changes in the carotid arteries of patients treated with unilateral neck radiation therapy (RT) for head and neck cancer.

**METHODS:** Bilateral carotid artery duplex studies were performed at 0, 3, 6, 12, 18 months and 2, 3, 4, and 5 years following RT. Intima media thickness (IMT); global and regional circumferential, as well as radial strain, arterial elasticity, stiffness, and distensibility were calculated.

**RESULTS:** Thirty-eight patients were included. A significant difference in the IMT from baseline between irradiated and unirradiated carotid arteries was detected at 18 months (median, 0.073mm vs -0.003mm; *P*=0.014), which increased at 3 and 4 years (0.128mm vs 0.013mm, *P*=0.016, and 0.177mm vs 0.023mm, *P*=0.0002, respectively). A > 0.073mm increase at 18 months was significantly more common in patients who received concurrent chemotherapy (67% vs 25%; *P*=0.03). A significant transient change was noted in global circumferential strain between the irradiated and unirradiated arteries at 6 months (median difference, -0.89, *P*=0.023), which did not persist. No significant differences were detected in the other measures of elasticity, stiffness, and distensibility.

**CONCLUSIONS:** Functional and morphologic changes of the carotid arteries detected by carotid ultrasound, such as changes in global circumferential strain at 6 months and carotid IMT at 18 months, may be useful for the early detection of radiation-induced carotid artery injury, can guide future research aiming to mitigate carotid artery stenosis, and should be considered for clinical surveillance survivorship recommendations after head and neck RT.

## INTRODUCTION

Radiation therapy (RT) is an integral part of the treatment of head and neck cancer.^1^ Most patients require treatment to the cervical lymph nodes adjacent to the carotid arteries, which most often cannot be excluded from the RT volume. This increases the risk for accelerated atherosclerosis, carotid artery stenosis and subsequent transient ischemic attack (TIA) and stroke.^2,3^ The long latent interval of several years from RT to the development of carotid artery stenosis makes it difficult to identify patients who will develop adverse outcomes.^4,5^

RT has been shown to be associated with morphologic changes in the carotid artery, in the form of increased wall thickness and accelerated atherosclerosis.^6^ Carotid intimal-medial thickness (IMT) is a measurement of the thickness of the artery wall using B-mode ultrasound. Increased common carotid artery (CCA) IMT is an early marker of atherosclerosis and a strong predictor of future vascular events in the general population.^7–9^ In patients exposed to head and neck RT, IMT has been shown to correlate well with the degree of subsequent carotid stenosis.^10–14^ The earliest timepoint that significant changes in IMT can be identified after RT has not been well studied. Furthermore, RT has also been shown to be associated with early vascular physiologic changes, including altered arterial elasticity. However, altered vascular elasticity is difficult to assess clinically. Imaging of vascular deformation with speckle tracking software, also known as strain, was shown in prior studies to correlate well with vascular elasticity parameters and has been suggested as a good surrogate for the assessment of carotid artery elasticity properties.^15–17^ However, serial changes in regional or global circumferential and radial strain, as well as changes in measures of elasticity, stiffness, and distensibility following RT of the neck, have not been prospectively evaluated. These measures might be able to detect early radiation-induced changes and identify patients who will benefit from preventive therapies.

Although several clinical risk factors have been associated with an increased risk of atherosclerosis, including age, hypertension, diabetes mellitus, hypercholesterolemia, and smoking,^18^ the effect of these factors on the progression of radiation-induced carotid artery disease, as well as the effect of risk factor–modifying therapies such as statins, aspirin, and antihypertensive and antidiabetic treatments, need further evaluation. Furthermore, there is weak evidence that IMT increases with higher doses of RT to the neck, suggesting a dose effect.^14,19^ However, retrospective study designs, small sample sizes, and short intervals from RT make it difficult to draw firm conclusions.

In this study, we prospectively evaluated longitudinal changes in IMT, strain, and other measures of elasticity, stiffness, and distensibility in the irradiated carotid artery compared to the unirradiated side over predefined follow-up timepoints in patients with head and neck cancer treated with ≥ 50 Gy unilateral neck RT. In addition, we assessed the effect of clinical parameters and preventive therapies on the development of increased IMT, atherosclerosis, carotid stenosis, TIA/stroke, or need for revascularization.

## METHODS

### PATIENT POPULATION

We identified all adult patients (age ≥ 18 years) with histologically confirmed cancer of the head and neck who were scheduled to undergo ≥ 50 Gy RT to 1 side of the neck of at The University of Texas MD Anderson Cancer Center between February 2015 and February 2017. Patients with a history of carotid endarterectomy or carotid angioplasty and stenting were excluded. A history of or planned neck dissection on the irradiated or unirradiated side was not an exclusion criterion. Eligible patients who were able to provide written informed consent were prospectively enrolled in the study. The study was approved by the Institutional Research Board of MD Anderson (ClinicalTrials.gov Identifier: NCT02069964).

### BASELINE CHARACTERISTICS

Demographic and clinical characteristics were prospectively collected prior to patients’ first RT, including age, sex, race, BMI, medication history, cancer treatment history, blood pressure, and basic laboratory values (hemoglobin, glucose, creatinine, liver, and lipid profile).

### CAROTID DUPLEX

Bilateral carotid artery duplex studies were performed by trained technicians at baseline and at predefined follow-up timepoints: 3, 6, 12, and 18 months and 2, 3, 4, and 5 years following the initial RT. 2D and Doppler images of bilateral carotid arteries were captured and interpreted following the Society of Radiologists in Ultrasound consensus conference guidelines.^20^ The CCA, internal carotid artery, and external carotid arteries on both sides of the neck were examined with the patient supine on an examination couch. The carotid waveforms and peak systolic and end diastolic velocities were recorded for the internal carotid artery and CCA at a Doppler angle of < 60°. The diagnostic criteria for internal carotid artery stenosis were based on peak systolic and end diastolic velocities, as well as internal carotid artery/CCA ratios.^20^ All images were captured using Philips echocardiography machines.

IMT was determined offline using a semi-automated technique, with far wall measurements taken at a B-mode longitudinal view of the posterior wall of the CCA, 2 cm proximal from the carotid bifurcation and away from any atherosclerotic plaques; the results were averaged over 3 readings on a magnified image (Figure 1). Two-dimensional long- and short-axis gray-scale cine loops of the CCA were acquired for speckle tracking and strain analysis. An offline strain analysis was performed using a workstation equipped with 2-D strain software (EchoPac 7.0, GE Vingmed Ultrasound). The software enables automatic identification of speckles in the vessel wall and subsequent tracking of their movement during the cardiac cycle. Global and peak circumferential as well as radial peak systolic strain were measured in both carotid arteries for each patient (Figure 2). Peak systolic and diastolic velocities, plaque size estimates, stenosis severity in bilateral common and internal carotid arteries, carotid intraluminal and adventitia-to-adventitia diameter, maximal systolic diameter, and minimal diastolic lumen diameter in M-mode were recorded, in addition to systolic and diastolic pressure. Measures of arterial elasticity, stiffness, and distensibility were calculated using validated formulas, outlined below:

**FIGURE 1.**
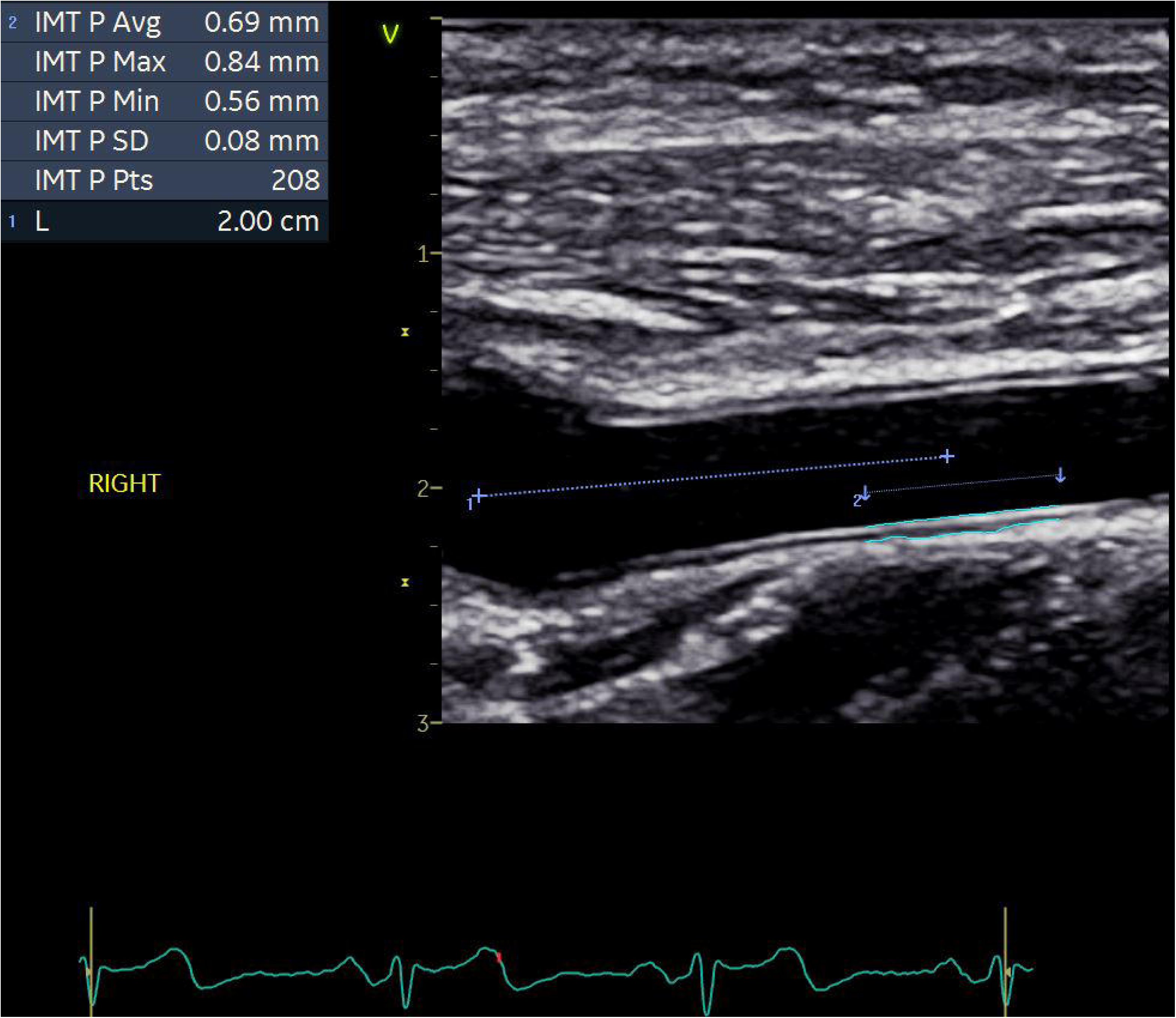
Intima Media Thickness measurements at the longitudinal view of the posterior wall of the CCA, 2 cm proximal from the carotid bifurcation and away from any atherosclerotic plaques; the results were averaged over 3 readings on a magnified image.

**FIGURE 2.**
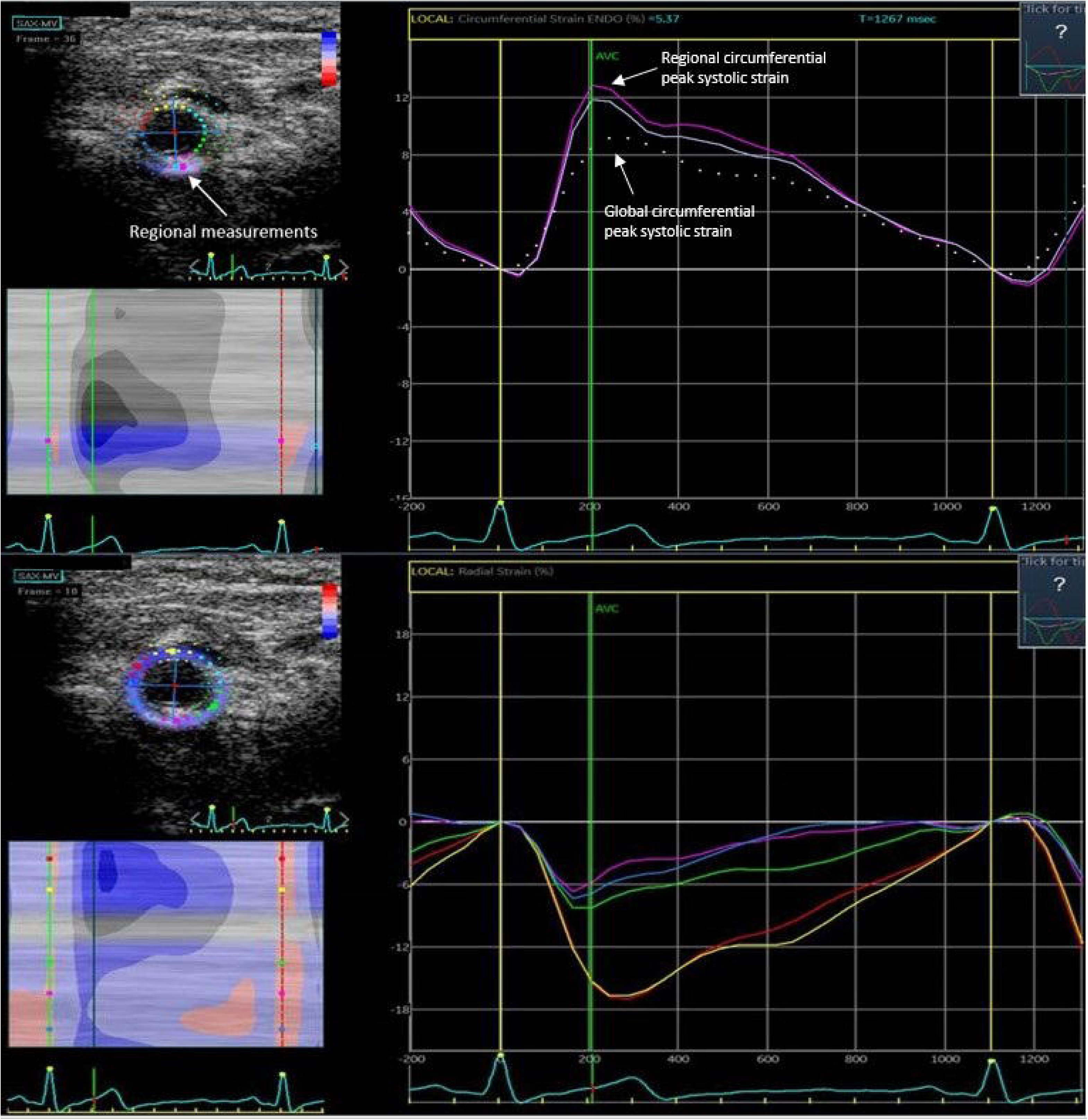
Measurement of circumferential global (dotted line) and regional (solid line) peak systolic strain (top) as well as radial peak systolic strain (bottom) using the speckle-tracking based strain analysis of the EchoPac 7.0, GE Vingmed Ultrasound

- Young’s modulus of elasticity (Y):^21^

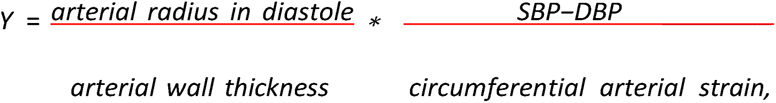

where SBP: systolic blood pressure and DBP: diastolic blood pressure

- Pressure-strain Young’s elastic modulus (E):^22^

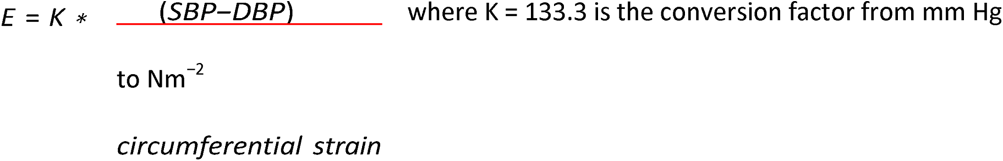

- Stiffness (stress-to-strain ratio) (β):^23^

β = ln (SBP/DBP)/circumferential strain

- Distensibility (1/β), adjusted to IMT:^22^

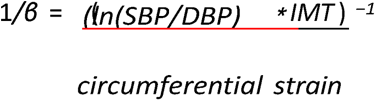

All studies were interpreted by a cardiologist expert in carotid duplex interpretation and was blinded to the patient’s medical history and treatment.

### ENDPOINTS

The primary endpoint of the study was the change in carotid IMT after RT between irradiated and unirradiated carotid arteries at predefined follow-up timepoints. Secondary endpoints included i. the change in carotid arterial wall strain (global or peak circumferential and radial) and other measures of arterial elasticity, stiffness, and distensibility following RT between irradiated and unirradiated carotid arteries at the predefined follow up-timepoints; ii. the incidence of atherosclerotic plaques, ≥ 50% carotid artery stenosis, TIA/stroke, or need for revascularization in irradiated carotid arteries compared to in unirradiated carotid arteries; and iii. the association between baseline clinical characteristics and risk-modifying therapies (HMGCoA reductase inhibitors, aspirin, and anti-hypertensive and -diabetic medications) and the IMT change in the irradiated and unirradiated carotid arteries.

### STATISTICAL ANALYSIS

The sample size was originally estimated to be 45 patients based on the results of a study by Dorresteijn et al (using a paired t test with mean difference of 0.3, standard deviation of 0.7, significance level (α) of 0.05 and 80% power).^13^ Although we had aimed to recruit a total of 60 patients to account for dropouts, relapses, and requirement for treatment to the unirradiated side, unexpected issues related to COVID-19 pandemic allowed us to enroll only 46 patients, out of which 38 had at least one follow up and were included in the analysis. Continuous variables were summarized as mean values +/- standard deviation (SD) or median values and interquartile ranges. Categorical variables are presented as frequencies and percentages. Categorical variables were compared between groups using Fisher’s exact test, and continuous variables were compared using the Wilcoxon rank-sum test.

The intima-media thickness (IMT) at 3, 6, 12, and 18 months and 2, 3, 4, and 5 years was compared to the baseline and the difference between the 2 was calculated for the irradiated and unirradiated arteries in each patient. The IMT change of the irradiated carotid artery was compared with the IMT change of the unirradiated carotid artery using a Wilcoxon rank-sum test at the predefined follow-up time points. The same test was also used to compare changes in strain and other measures of elasticity, stiffness, and distensibility from baseline between irradiated and unirradiated carotid arteries. A generalized linear mixed-effects model analysis was used to assess the association between clinical parameters and changes in IMT between irradiated and unirradiated carotid arteries over time. All statistical analyses were performed using R version 4.2.2 at a significance level of 5%. No adjustments for multiple testing were made.

## RESULTS

A total of 46 patients were enrolled in the study, 38 of whom were included in the analysis (5 were lost in follow up, 1 had poor quality of baseline images, 1 had significant undiagnosed carotid artery disease and 1 had history of carotid endarterectomy discovered after enrollment; Figure 3). The mean patient age was 59.1 ± 12.5 years; 40% of patients were female, and all were white. The mean BMI was 30.4 ± 6.1 kg/m^2^. Former or current tobacco use was reported in 43% of patients, while 34% of patients were treated for hypertension, 11% for diabetes mellitus; statins were used by 37% and aspirin by 29% of patients. Twenty one percent of patients did not have lymph node involvement and were treated with 50-56Gy of RT, while 79% had lymph node involvement requiring a higher RT dose (66-70Gy). RT was delivered to the left side of the neck in 70% and the right side in 30% of patients. The mean total radiation dose was 62.8 ± 3.7 Gy, delivered at a mean of 30.7 ± 1.4 fractions. The maximum dose delivered to the carotid artery was 63.4 ± 5.6 Gy, and the mean dose was 58.1 ± 4.2 Gy. Eight percent of patients received induction and 39% concurrent chemotherapy. Additional demographic and clinical characteristics are presented in Table 1. Serial ultrasonographic evaluations were performed, with 95% of patients completing the 3- and 6-month follow up evaluation, 87% the 12-month, 89% the 18-month, 84% the 2-year, 76% the 3-year, 55% the 4-year, and 18% the 5-year (the 4- and 5-year follow-up were mostly impacted by COVID-19 restrictions; Figure 3).

**FIGURE 3.**
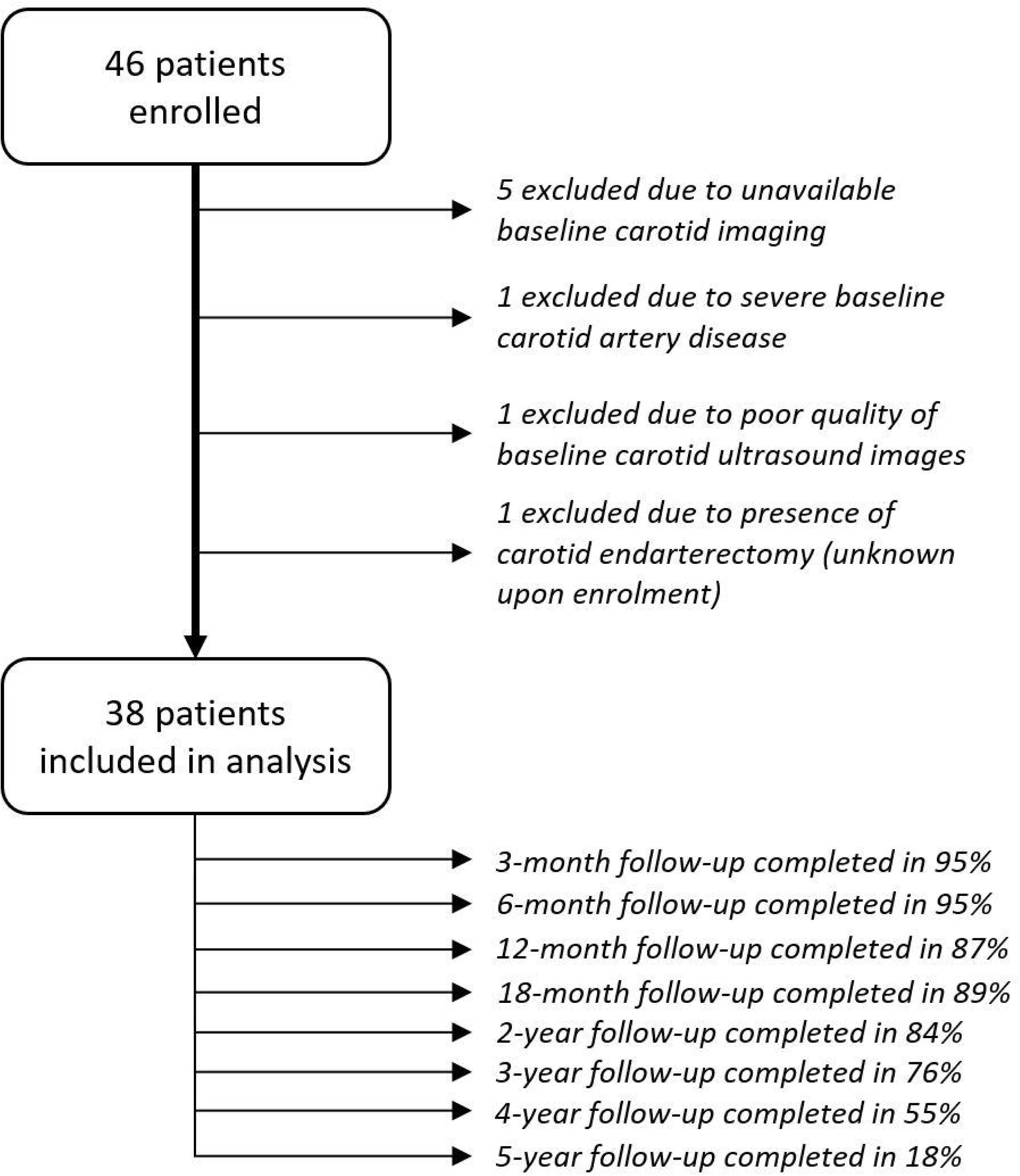
Number of Patients Who Were Enrolled in the Study, Included in the Analysis, and Completed Follow-Up

**TABLE 1.**
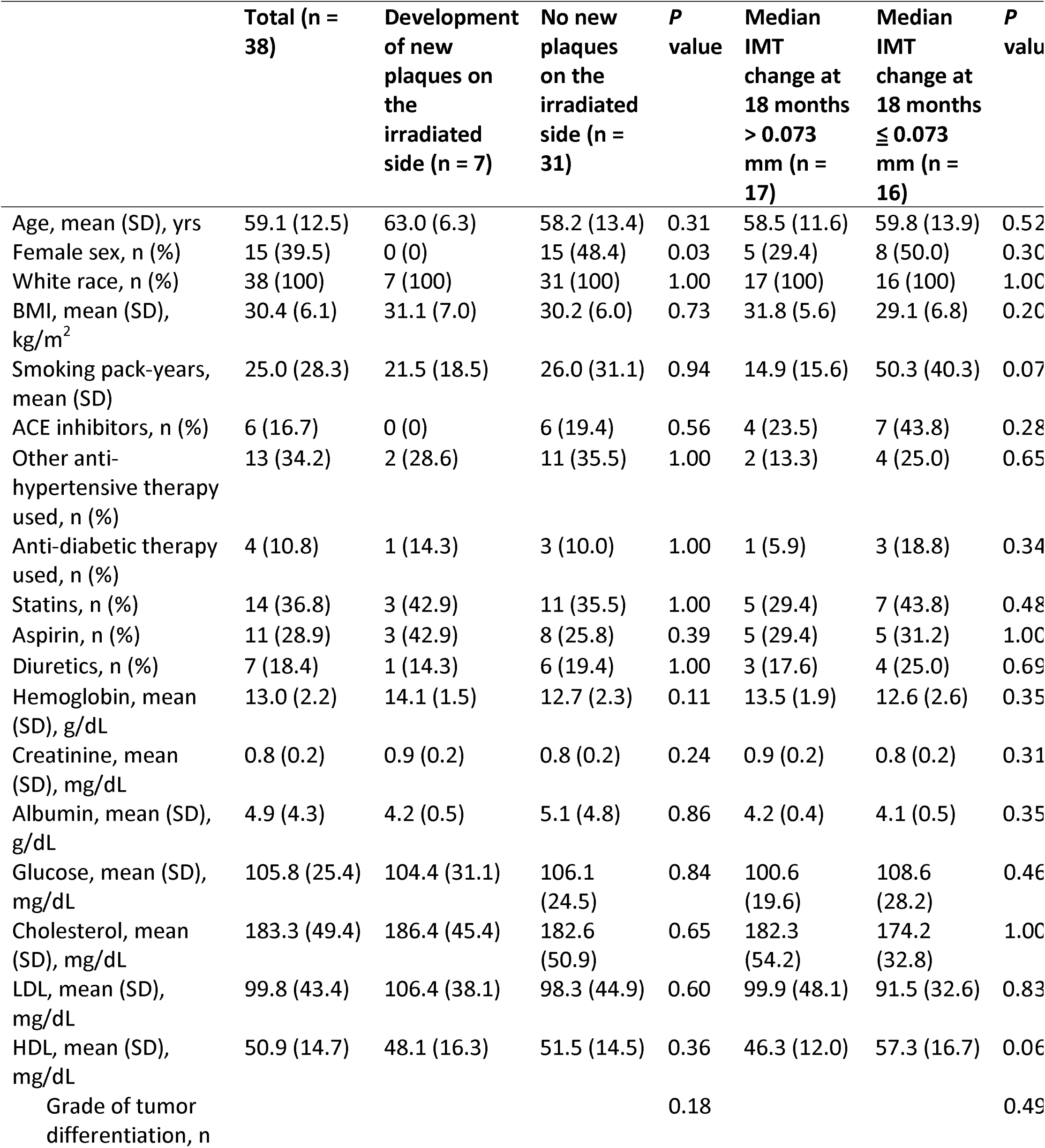
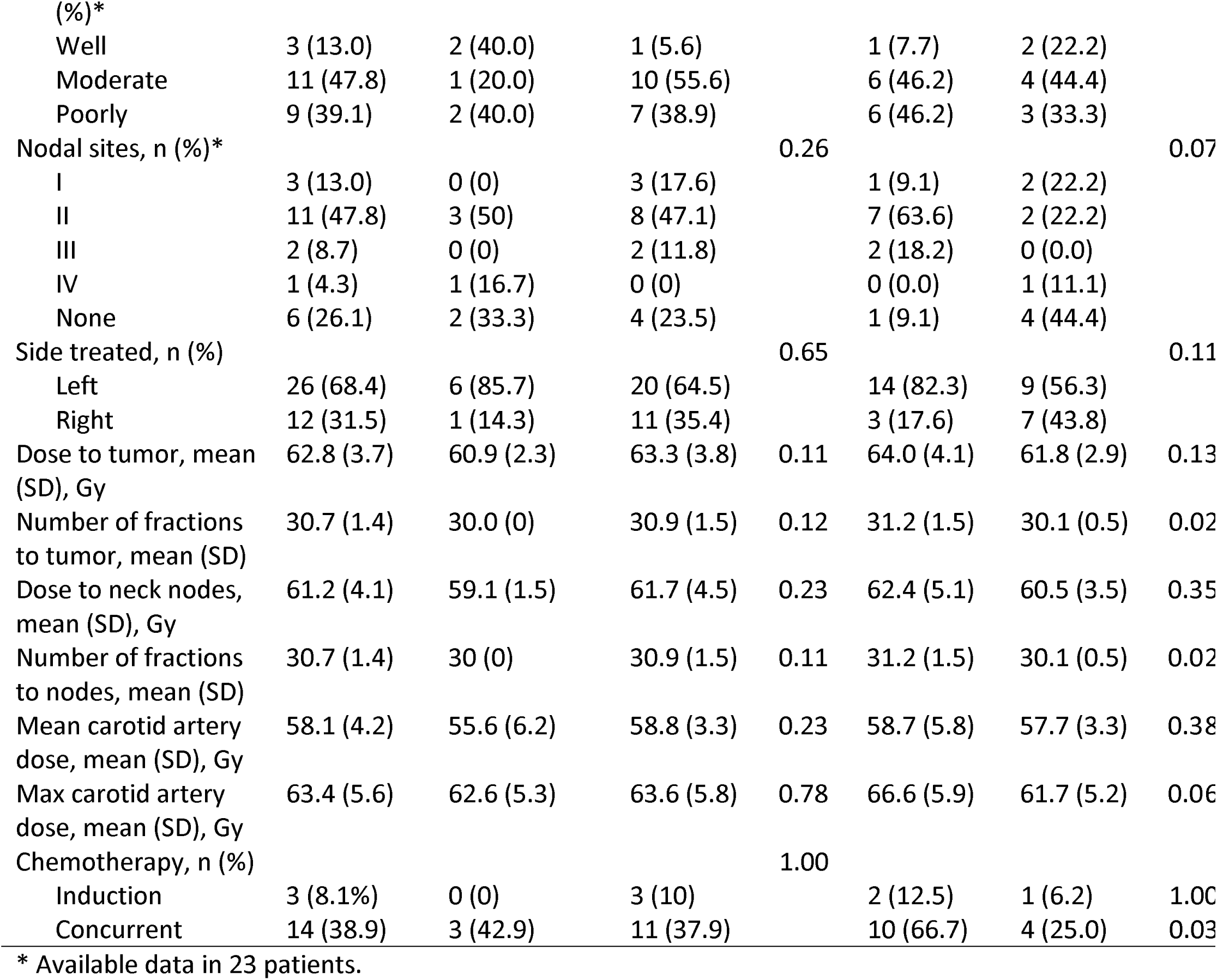
Demographic and Clinical Characteristics of Patients Based on Whether They Developed the Composite Outcome of Development of Atherosclerotic Plaques, ≥ 50% Stenosis, TIA/Stroke, or Need for Revascularization in the Irradiated Carotid Artery

### ULTRASONOGRAPHIC FINDINGS: IMT

A significant change in IMT from baseline between the irradiated and unirradiated carotid arteries was detected as early as 18 months from the date of first RT (median change, 0.073 mm vs -0.003 mm; *P* = 0.014). Although the median IMT value of the irradiated carotid arteries in the entire cohort was statistically higher than that of the unirradiated side even at 12 months (0.79 vs 0.73 mm; *P* = 0.033), the IMT change for each individual patient from baseline significantly differed only at 18 months. The difference remained significant and increased at 3 and 4 years (0.128 mm vs 0.013 mm, *P* = 0.016, and 1.777 mm vs 0.023 mm, *P* = 0.0002, respectively; Table 2, Figure 4). Patients with an IMT change > 0.073 mm in the irradiated carotid artery were more likely to have received concurrent chemotherapy than were those with an IMT change ≤ 0.073 mm at 18 months (median value) (67% vs 25%; *P* = 0.03; Table 1). In addition, patients with an IMT change > 0.073 mm at 18 months received a higher number of radiation fractions to the tumor and neck lymph nodes than did those with an IMT change of ≤ 0.073 mm (31.2 [1.5] vs 30.1 [0.5]; *P* = 0.05).

**FIGURE 4.**
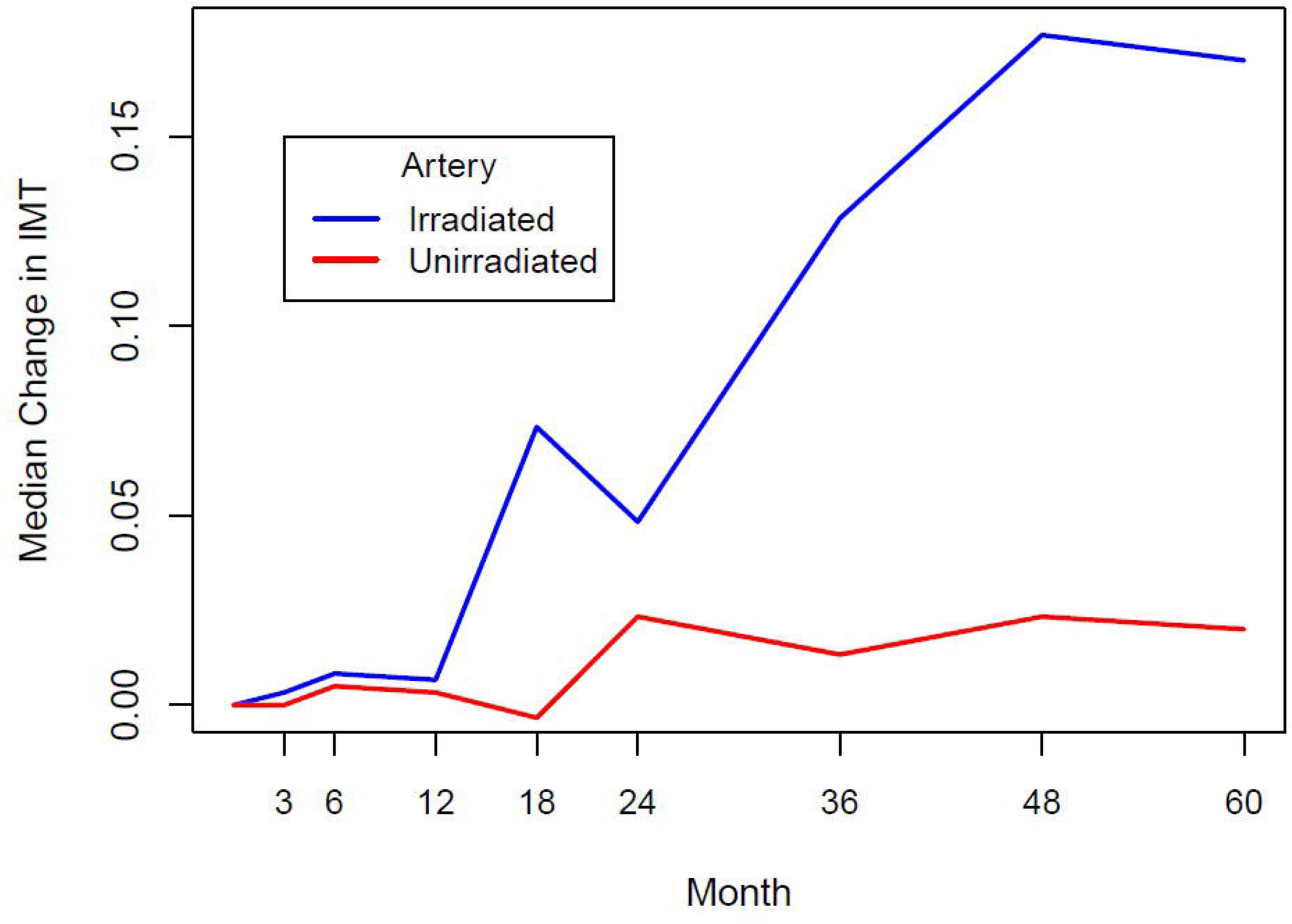
Median Change in Intima-Media Thickness (IMT) from Baseline Over Time Between Irradiated and Unirradiated Carotid Arteries

**TABLE 2.** Median Intima-Media Thickness (IMT) Change from Baseline Between Unirradiated and Irradiated Carotid Arteries; Differences Were Significant at 18 Months and Later.

| **Time** | **No. of patients** | **Median change:**<br>**unirradiated artery** | **Median change:**<br>**irradiated artery** | ***P* value** |
| --- | --- | --- | --- | --- |
| 3 months | 36 | 0.000 | 0.003 | 0.84 |
| 6 months | 36 | 0.005 | 0.008 | 0.72 |
| 12 months | 33 | 0.003 | 0.007 | 0.88 |
| 18 months | 34 | -0.003 | 0.073 | 0.014 |
| 2 years | 32 | 0.023 | 0.048 | 0.013 |
| 3 years | 29 | 0.013 | 0.128 | 0.016 |
| 4 years | 21 | 0.023 | 0.177 | 0.0002 |
| 5 years | 7 | 0.020 | 0.17 | 0.078 |

In a linear mixed-effect regression analysis evaluating the association of several demographic and clinical parameters with changes in IMT over time, there was a trend towards a significant association between aspirin (0.12 [-0.01-0.25]; *P* = 0.08) and statin use (0.11 [- 0.004-0.23]; *P* = 0.07), as well as between lower mean radiation dose to the carotid artery (0.008 [-0.001-0.03]; *P* = 0.08) and a smaller IMT change.

### DIFFERENCE IN PEAK STRAIN, ELASTICITY, STIFFNESS, DISTENSIBILITY, AND COMPLIANCE OF IRRADIATED AND UNIRRADIATED CAROTID ARTERIES OVER TIME

Changes from baseline in the peak radial, as well as peak and global circumferential strains for the irradiated and unirradiated carotid arteries over time are presented in Table 3. A significant difference was noted in the change in the global circumferential strain from baseline between the irradiated and unirradiated carotid arteries at 6 months (median difference = -0.89, *P* = 0.023), which did not persist in follow-up studies. The rest of the measurements were not different between the 2 groups.

**TABLE 3.** Comparison of Differences in Peak Radial, Peak Circumferential, and Global Circumferential Strain Between Irradiated and Unirradiated Carotid Arteries Over Time.

| **Time** | **Measurement** | **Median change: irradiated (mm)** | **Median change: unirradiated (mm)** | **Median difference (mm)** | **P value** |
| --- | --- | --- | --- | --- | --- |
| 3 months | Peak Radial | -0.16 | 2.18 | -0.12 | 0.23 |
|  | Peak Circumferential | -0.52 | -0.91 | 1.34 | 0.61 |
|  | Global Circumferential | -0.12 | 0.90 | -0.68 | 0.37 |
| 6 months | Peak Radial | -0.63 | 2.03 | -4.05 | 0.062 |
|  | Peak Circumferential | 0.55 | 1.56 | -3.10 | 0.32 |
|  | Global Circumferential | -0.31 | 0.99 | -0.89 | 0.023 |
| 12 months | Peak Radial | -0.53 | 3.54 | -4.53 | 0.09 |
|  | Peak Circumferential | 0.37 | -0.21 | -4.37 | 0.35 |
|  | Global Circumferential | -0.56 | -0.47 | -1.09 | 0.40 |
| 18 months | Peak Radial | 0.80 | 2.65 | -1.60 | 0.43 |
|  | Peak Circumferential | 0.95 | 0.75 | -1.01 | 0.75 |
|  | Global Circumferential | -0.33 | -0.40 | -0.53 | 0.46 |
| 2 years | Peak Radial | 0.03 | 3.95 | -2.99 | 0.065 |
|  | Peak Circumferential | -0.40 | 0.04 | -0.16 | 0.92 |
|  | Global Circumferential | -0.22 | -0.29 | -0.76 | 0.17 |
| 3 years | Peak Radial | 1.48 | 2.90 | -3.16 | 0.41 |
|  | Peak Circumferential | -3.04 | -2.40 | 0.41 | 0.78 |
|  | Global Circumferential | -0.43 | -1.00 | -0.66 | 0.29 |
| 4 years | Peak Radial | 0.18 | 4.11 | -1.76 | 0.14 |
|  | Peak Circumferential | -1.07 | 0.55 | -1.61 | 0.52 |
|  | Global Circumferential | 0.37 | 0.80 | -0.12 | 0.89 |
| 5 years | Peak Radial | 4.15 | -5.18 | 3.58 | 0.47 |
|  | Peak Circumferential | -0.19 | 1.87 | -6.22 | 0.16 |
|  | Global Circumferential | -2.24 | -4.46 | 0.93 | 0.38 |

Changes in elasticity, stiffness, and distensibility between the irradiated and unirradiated arteries over time are presented in Table 4. No significant differences were detected between the 2 sides.

**TABLE 4.** Changes in Elasticity, Stiffness, and Distensibility Between Irradiated and Unirradiated Arteries Over Time.

| **Time** | **Measurement** | **Median change: irradiated** | **Median change: unirradiated** | **Median difference** | **P value** |
| --- | --- | --- | --- | --- | --- |
| 3 months | Y: Young's Modulus | 1.28 | 1.51 | -16.27 | 0.97 |
|  | E: Pressure-strain Young's Modulus | -126.99 | 736.93 | -1377.81 | 0.13 |
|  | Beta: Stiffness | -0.01 | 0.06 | -0.11 | 0.10 |
|  | Distensibility | 0.63 | -0.26 | 1.52 | 0.60 |
| 6 months | Y: Young's Modulus | 2.16 | 2.04 | 18.68 | 0.075 |
|  | E: Pressure-strain Young's Modulus | -184.83 | 1106.21 | -131.65 | 0.80 |
|  | Beta: Stiffness | 0.02 | 0.06 | -0.01 | 0.87 |
|  | Distensibility | 1.19 | 2.43 | -0.79 | 0.36 |
| 12 months | Y: Young's Modulus | 11.25 | 10.43 | 12.20 | 0.43 |
|  | E: Pressure-strain Young's Modulus | 399.15 | 522.87 | 73.49 | 0.81 |
|  | Beta: Stiffness | 0.02 | 0.04 | 0.02 | 0.64 |
|  | Distensibility | -0.42 | 1.67 | -2.04 | 0.36 |
| 18 months | Y: Young's Modulus | 7.11 | 8.22 | -5.05 | 0.28 |
|  | E: Pressure-strain Young's Modulus | 70.71 | 1085.26 | -2109.36 | 0.89 |
|  | Beta: Stiffness | 0.01 | 0.11 | -0.25 | 0.80 |
|  | Distensibility | 1.42 | -0.37 | 5.55 | 1.00 |
| 2 years | Y: Young's Modulus | 26.58 | 5.17 | 23.01 | 0.11 |
|  | E: Pressure-strain Young's Modulus | 1059.41 | 82.22 | 1670.39 | 0.23 |
|  | Beta: Stiffness | 0.09 | 0.03 | 0.12 | 0.25 |
|  | Distensibility | -0.31 | 0.59 | -0.56 | 0.71 |
| 3 years | Y: Young's Modulus | 37.18 | 1.81 | 18.67 | 0.41 |
|  | E: Pressure-strain Young's Modulus | 409.50 | 268.15 | -2521.25 | 0.98 |
|  | Beta: Stiffness | -0.05 | 0.02 | -0.18 | 1.00 |
|  | Distensibility | -4.65 | -0.62 | -0.27 | 0.58 |
| 4 years | Y: Young's Modulus | 11.97 | 26.37 | 13.65 | 0.52 |
|  | E: Pressure-strain Young's Modulus | 2308.17 | 1735.57 | 953.34 | 0.37 |
|  | Beta: Stiffness | 0.15 | 0.11 | 0.07 | 0.31 |
|  | Distensibility | -1.08 | 2.09 | -2.88 | 0.37 |
| 5 years | Y: Young's Modulus | 24.79 | -2.49 | -0.31 | 0.81 |
|  | E: Pressure-strain Young's Modulus | 1849.92 | -145.96 | 200.88 | 0.69 |
|  | Beta: Stiffness | 0.11 | -0.01 | -0.01 | 0.81 |
|  | Distensibility | -2.10 | 2.14 | -9.12 | 0.22 |

### DEVELOPMENT OF ATHEROSCLEROTIC PLAQUES, CAROTID STENOSIS, TIA/STROKE, OR NEED FOR REVASCULARIZATION

At baseline, 7 patients (18%) had evidence of atherosclerotic plaques on the side that was subsequently irradiated and 8 (21%) on the side that was not subsequently irradiated. Seven patients developed evidence of new atherosclerotic plaques on the irradiated side at a median of 12 months, compared to 5 on the unirradiated side at a median of 18 months (*P* = 0.74; 3 of these patients developed new atherosclerotic plaques on both the irradiated and unirradiated sides). All 7 patients had developed evidence of new atherosclerotic plaque by the second year of follow up (7/32 patients, 22%). Of these, 2 patients developed ≥ 50% carotid stenosis on the irradiated side at 1 year follow up (2/33, 6%) vs 1 patient on the unirradiated side at 4 years. The patients who developed ≥ 50% stenosis had no atherosclerotic plaques at baseline. One patient (3%) out of those with ≥ 50% stenosis, experienced a stroke on the side of the irradiated carotid artery, and was treated with revascularization and medical therapy. No patients experienced a TIA or stroke on the side of the unirradiated carotid artery. All patients who developed new atherosclerotic plaques, with or without ≥ 50% stenosis on the irradiated side were male compared to the patients who did not develop new atherosclerotic plaques on the same side (100% vs 51.6%; *P*=0.03). Otherwise, no significant differences in the demographic and clinical characteristics were noted between the 2 groups (Table 1).

## DISCUSSION

This prospective study evaluated longitudinal changes in carotid artery morphology and functional characteristics using traditional and novel techniques (carotid IMT, carotid arterial wall stain, and other measures of elasticity, stiffness, and distensibility) at baseline and following unilateral neck irradiation for head and neck cancer. We found significant changes in carotid IMT as early as 18 months after unilateral neck RT in the irradiated compared to the contralateral, unirradiated carotid artery. Concurrent chemotherapy was associated with a larger increase in IMT at 18 months. Furthermore, significant changes in global circumferential strain were detected 6 months after RT. One of 5 patients developed new atherosclerotic plaques in the irradiated carotid artery at a median of 2 years.

The traditionally expected latent period separating initial radiation-induced vascular injury during cancer treatment and the late stage of development of clinical cardiovascular complications makes the investigation of radiation-induced vascular diseases challenging. In our study, 2 patients who had no atherosclerotic plaque at baseline, developed ≥ 50% carotid stenosis on the irradiated side vs. 1 patient on the unirradiated side, suggesting that some patients receiving RT may be at risk for accelerated atherogenesis, highlighting the need for earlier screening. Identifying simple diagnostic tools that detect radiation-related carotid changes early will help with patient risk stratification and subsequent investigation the identification of therapies that mitigate radiation-induced cardiovascular disease. Mitigation of radiation-induced carotid artery disease is an important aspect of the care of head and neck cancer survivors and highlights the significance of the findings of this study.

RT has been shown to be associated with early morphologic vascular changes, such as increased IMT, and vascular physiologic changes, such as altered arterial elasticity.^6^ A recent systematic review by Randolph et al. identified a total of 8 studies published after 2010, 4 of which were prospective, that evaluated carotid IMT change after RT.^24^ All 8 studies showed significant increases in carotid IMT after neck irradiation.^24^ Three of the 4 prospective studies, included patients who underwent bilateral neck irradiation and reported a significant IMT increase from baseline as early as 6 weeks after RT (sample size, 19-50 patients).^25–27^ The fourth prospective study by Wilbers et al. compared IMT changes between irradiated and unirradiated carotid arteries among 48 patients and reported a 5 times larger increase in IMT in the irradiated carotid artery at a median follow-up of 6.7 years.^28^ Compared to the above-mentioned prospective studies, ours is the first that longitudinally compared IMT changes in the irradiated versus the unirradiated carotid arteries at multiple, frequent time points in patients who underwent unilateral neck RT. Compared to previous studies that used patients without cancer as control, our study used the contralateral, unirradiated carotid artery to control for baseline comorbidities and confounders that might have biased the results when patients with cancer were compared to patients without cancer. Furthermore, compared to previous studies with a single follow-up or a few short-term follow-up time points, our study included multiple serial assessments (from 3 months to 5 years after RT). We found a significant IMT increase in the irradiated compared to the unirradiated carotid artery as early as 18 months after RT, which persisted at 3 and 4 years. At 5 years, the IMT change from baseline was 7.5 times higher in the irradiated carotid artery than in the unirradiated artery, which is in line with the results of the study by Wilbers et al.^28^ In our cohort, no significant IMT changes from baseline were noted in the irradiated versus the unirradiated carotid arteries at 3, 6, or 12 months after RT.

Several clinical risk factors have been associated with increased risk of atherosclerosis, including age, hypertension, diabetes mellitus, hypercholesterolemia, and smoking;^18^ however, the effect of these factors and of risk factor–modifying therapies, such as statins, aspirin, and antihypertensive and antidiabetic treatments, in radiation-induced carotid artery disease progression has not been well evaluated. Furthermore, there is weak evidence that IMT increases with higher doses of RT to the neck, suggesting a dose effect.^14,19^ In our study, we prospectively evaluated patients with head and neck cancer who had been treated with RT and were followed up at frequent time intervals and showed that patients with larger carotid IMT changes (> 0.073 mm) were more frequently treated with concurrent chemotherapy. No significant differences in age or baseline cardiovascular comorbidities were detected between patients with a larger (> 0.073 mm at 18 months) vs smaller IMT change (≤ 0.073 mm at 18 months). Furthermore, there was a trend towards a statistically significant difference between RT dose and IMT change and a statistically but not clinically significant difference between the number of radiation fractions and IMT change. Of note, RT doses given were within a relatively narrow range, potentially obfuscating a dose-response effect. The absence of an association between age, baseline cardiovascular comorbidities, or radiation dose and larger IMT change might be related to the small sample size in the study. The results of our analysis also showed a trend towards a statistically significant association between aspirin and statin use and a smaller IMT change, suggesting that aspirin and statin have a potential beneficial role in mitigating IMT increase after RT. However, these results should be interpreted with caution given the number of variables analyzed.

Other than the early morphologic vascular changes, RT has also been associated with vascular physiologic changes, including altered arterial elasticity.^6^ Although altered vascular elasticity is difficult to assess clinically, previous studies have shown a good correlation between traditionally measured vascular elasticity parameters and vascular strain and have proposed the use of vascular strain for the assessment of carotid artery elasticity.^15,16^ Changes in carotid vascular tissue deformation (strain) after RT have not been well studied. Our prospective study evaluated contiguous changes in the global and peak circumferential, as well as radial strain of irradiated carotid arteries compared to the contralateral unirradiated carotid arteries in patients with head and neck cancer. A significant difference was noted in the global circumferential strain between the irradiated and unirradiated carotid arteries at 6 months (median difference = -0.89, *P* = 0.023), which did not persist in follow-up studies. This difference might be related to subacute transient inflammatory changes that affect vascular tissue deformation and subsequently resolve or stabilize. Global circumferential strain imaging may detect early elasticity changes in the setting of vascular injury. This finding needs to be further tested in larger studies. No significant changes were noted in the peak circumferential or radial strain of the irradiated versus the unirradiated carotid arteries over time.

Traditional measures of elasticity, stiffness, and distensibility, although laborious to obtain and reproduce, have been used in previous studies to assess functional changes after RT. A retrospective study by Gujral et al. assessed elasticity (elastic modulus) and stiffness (beta stiffness index [β]) in 50 patients with head and neck cancer, ≥ 2 years after unilateral neck RT; the results were compared with the changes to the contralateral, unirradiated carotid arteries. The authors reported significant changes in elastic modulus but no significant changes in the beta stiffness index.^29^ Our study prospectively evaluated changes in carotid artery elasticity, expressed by Young’s modulus of elasticity, pressure-strain Young’s Modulus equations, beta stiffness, and distensibility, adjusted to IMT (1/β) between the irradiated and unirradiated arteries over time, in patients with head and neck cancer. No significant differences were detected between the 2 sides over time, suggesting that these measures are not sensitive enough to detect early changes in radiation-related carotid artery disease.

Several studies have shown that patients treated with neck RT are at increased risk of developing carotid artery stenosis. In a recent meta-analysis of 19 studies, carotid stenosis of ≥ 50% was reported in 4% (95% CI: 2%-5%) of patients after 1 year, 12% (95% CI: 9%-15%) after 2 years, and 21% (95% CI: 9%-36%) after 3 years from RT. In our cohort, 22% of patients developed evidence of new atherosclerotic plaques at a median of 2 years and 6% developed ≥ 50% carotid stenosis at 1 year follow up. Importantly, all patients who developed ≥ 50% carotid stenosis in our cohort had the stenosis developed at 1 year follow up, with no additional patients developing stenosis in the years 2-4. All patients who developed ≥ 50% stenosis had no atherosclerotic plaques at baseline. This might indicate that there are two categories of patients who develop carotid artery disease following RT; the first category includes those with hyper-acute accelerated atherosclerosis with development of ≥ 50% stenosis within 1 year of follow up; and the second category includes patients with slower progression of atherosclerosis following RT.

The strengths of this study include its prospective nature, the thorough evaluation of the morphologic and functional characteristics of both the irradiated and unirradiated carotid arteries in the same patient, which minimizes the effect of confounders, as well as the frequent follow-up assessments. This study also has some limitations, the main one being the small sample size. Follow up was affected by the restrictions that the COVID-19 pandemic posed during the 4^th^ and 5^th^ year. Despite the small sample size, we were still able to obtain multiple serial ultrasonographic measurements and assess the morphology and function of the carotid arteries with enough statistical power to detect the findings reported above. All patients in our study were white, which did not allow for an assessment of racial differences, and patients were not followed up past 5 years after RT. This relatively brief follow-up period and the subsequent small number of clinical outcomes in our cohort (carotid stenosis, stroke/TIA, or need for revascularization) did not allow us to make correlations between the increased IMT noted at 18 months and the subsequent development of clinically significant stenosis.

In conclusion, alterations in the functional and morphologic characteristics of the carotid arteries following exposure to RT include significant early changes in global circumferential strain at 6 months and carotid IMT at 18 months. These 2 easy and simple-to-measure parameters may be useful tools for the early detection of radiation-induced carotid injury and can guide future research in assessing the impact of preventive therapies, including statins and modified RT regimens, to reduce the long-term complications of radiation-induced carotid artery stenosis.

## Data Availability

All data produced in the present study are available upon reasonable request to the authors

## Acknowledgements

The authors would like to acknowledge the team of Editing Services, Research Medical Library at MD Anderson Cancer Center for their help with editing the manuscript.

